# Psychophysical pain encoding in the cingulate cortex predicts responsiveness of electrical stimulation

**DOI:** 10.1101/2023.03.18.23287266

**Authors:** Rose M. Caston, Elliot H. Smith, Tyler S. Davis, Hargunbir Singh, Shervin Rahimpour, John D. Rolston

## Abstract

**Background:** The anterior cingulate cortex (ACC) plays an important role in the cognitive and emotional processing of pain. Prior studies have used deep brain stimulation (DBS) to treat chronic pain, but results have been inconsistent. This may be due to network adaptation over time and variable causes of chronic pain. Identifying patient-specific pain network features may be necessary to determine patient candidacy for DBS.

**Hypothesis:** Cingulate stimulation would increase patients’ hot pain thresholds if non-stimulation 70-150 Hz activity encoded psychophysical pain responses.

**Methods:** In this study, four patients who underwent intracranial monitoring for epilepsy monitoring participated in a pain task. They placed their hand on a device capable of eliciting thermal pain for five seconds and rated their pain. We used these results to determine the individual’s thermal pain threshold with and without electrical stimulation. Two different types of generalized linear mixed-effects models (GLME) were employed to assess the neural representations underlying binary and graded pain psychophysics.

**Results:** The pain threshold for each patient was determined from the psychometric probability density function. Two patients had a higher pain threshold with stimulation than without, while the other two patients had no difference. We also evaluated the relationship between neural activity and pain responses. We found that patients who responded to stimulation had specific time windows where high-frequency activity was associated with increased pain ratings.

**Conclusion:** Stimulation of cingulate regions with increased pain-related neural activity was more effective at modulating pain perception than stimulating non-responsive areas. Personalized evaluation of neural activity biomarkers could help identify the best target for stimulation and predict its effectiveness in future studies evaluating DBS.

## Introduction

Pain is a complex, multidimensional psychological process with neural activity spanning networks involved in sensory-discriminative, affective-evaluative, and cognitive-evaluative dimensions [1]. The cingulate cortex plays a crucial role in the affective and cognitive aspects of pain processing. Specifically, the anterior cingulate cortex (ACC) comprises multiple subregions supporting various functions related to emotion, motivation, higher cognition, and motor control [2–5]. The cingulate cortex has been a target of interest in neurosurgical pain treatment for over 60 years, largely via ablation [5–8].

More contemporary targeted neurosurgical treatment involves deep brain stimulation (DBS) of the ACC. Recent case reports in non-pain patients [9,10] and one patient with refractory neuropathic pain [11] showed promising results. A case series and prospective review found that only a handful of patients had adequate pain relief three years later [12–14]. In interpreting the state of ACC DBS after these studies and how to move forward, it is important to consider that the larger study population consisted of many types of chronic pain syndromes.

While there is some overlap between chronic pain etiologies, each patient’s cognitive and emotional factors are unique and can cause functional reorganization in their neural connectivity over time [15]. Identifying patient-specific pain network features that can explain and predict the natural fluctuations of sustained pain is necessary to identify structures and patients that will respond to DBS. We propose that ACC neural activation associated with ongoing pain can explain whether a patient will respond to pain modification using DBS. Pain researchers generally rely on inducing pain in healthy volunteers using brief, millisecond-long stimuli with infrared lasers [16–18]. Brief pain stimuli may be limited for studying cognitive and emotional factors’ involvement in painful experiences. However, experimental tonic pain is thought to be more liable to elicit sensory, emotional, and cognitive encoding-based processes like those experienced by chronic pain patients [1].

Our approach utilized intracranial neurophysiological recordings from four patients undergoing stereo-electroencephalography (sEEG) for seizure focus localization. Patients underwent a psychophysical pain task with and without electrical stimulation of the ACC. We assessed whether stimulation changed patients’ hot pain threshold. We also evaluated the relationship between non-stimulation behavioral pain responses and changes in broadband high-frequency local field potential amplitude (HFA; 70-150 Hz) [19–21]. Our findings demonstrate that patients who displayed increased pain-related neural activity responded to pain modulation through stimulation.

## Methods

### Psychophysical Pain Task

Our research group recently developed a thermoelectric device compatible with intracranial electrodes. The device and integrated software have been validated as a psychophysical pain task in healthy human subjects [22]. The task consists of four main events (**Figure 1A**), which occur in each trial: 1) The participant places their hand on the device for 5 seconds, 2) the participant removes their hand from the device, 3) the participant responds to whether the trial was perceived as painful, and 4) the participant rates the perceived pain and heat intensity on a scale of 0-10. Each of these events is time-locked to the ongoing intracranial recording. The start and end of each trial are time-locked using a capacitive touch sensor under the device’s surface. The patient used their dominant hand unless an intravenous access site was on that hand (Patient 1: right hand, Patient 2: left hand, Patient 3: right hand, Patient 4: left hand).

**Figure 1.**
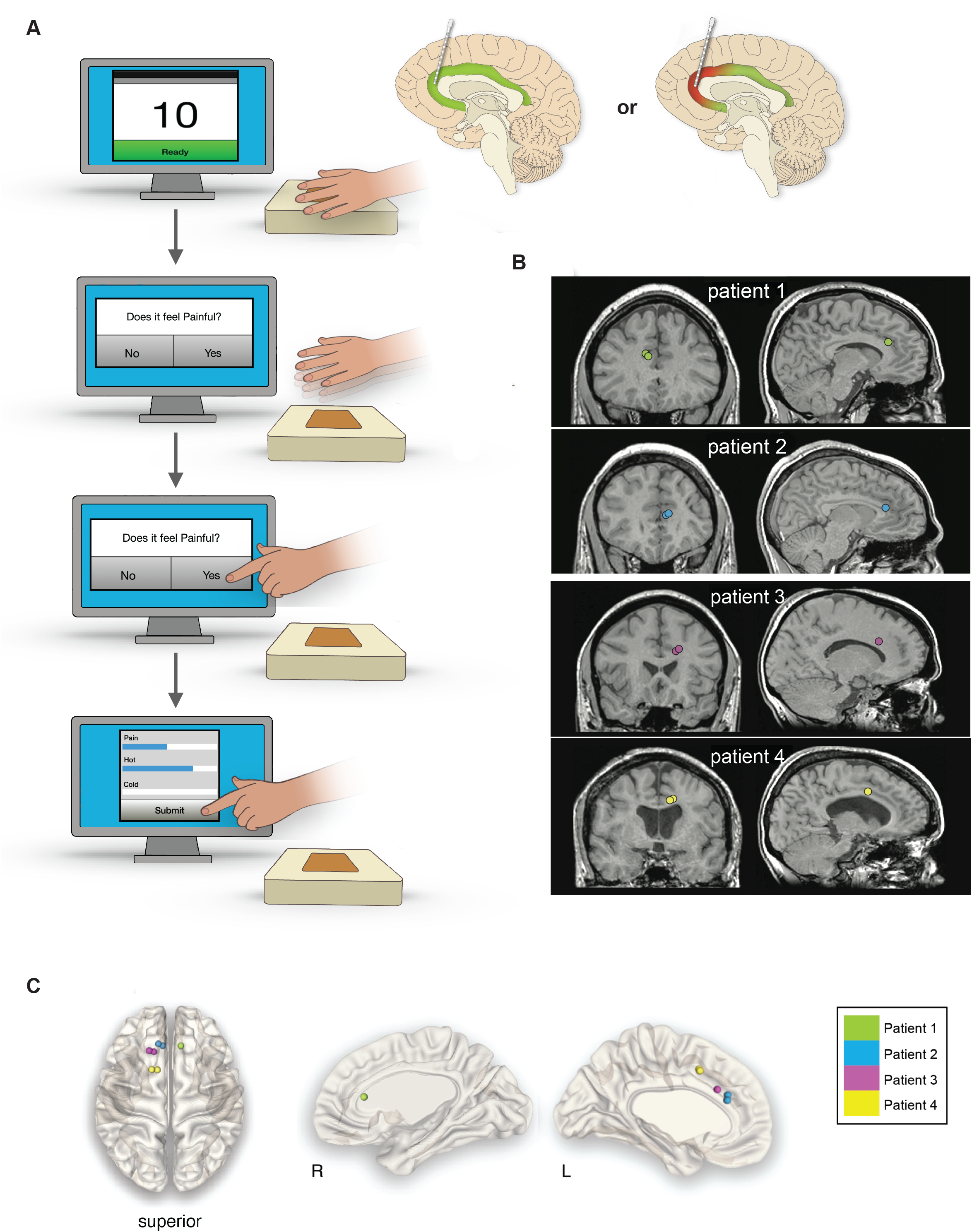
Psychophysical task paradigm and intracranial stimulation locations. 1A: Pain stimuli of varying intensity were applied to the hand’s palmar surface in patients with intracranial electrodes. The intensity of the stimulus was adjusted in each trial. One trial constituted having the hand on the stimulus for 10 seconds (top screen). Stimulation was applied in a pseudo-random order. The patient removed their hand from the device. After removing their hand, the patient responded to the question, “Was that painful?” on the touchscreen desktop. After answering “yes” or “no,” the patient filled out how painful, hot, or cold the stimulus felt on a scale of 0-10 (bottom right screen). After submitting their answers, the stimulus was adjusted, and the sequence was repeated. 1B: Four different patients underwent stimulation of their cingulate cortex. While many more electrodes were implanted in each patient’s brain, we only show the ones stimulated for a psychophysical effect. Electrode locations are displayed according to the coordinates in MNI space. Two spheres represent bipolar stimulation performed with Patient 2, 3, and 4. Monopolar stimulation was performed in Patient 1. 1C: Location of stimulation electrodes overlayed on patients’ preoperative MRI and postoperative CT. Electrode locations were visualized in the LeGUI software, following the method in *Electrodes*.

The thermoelectric device also includes built-in software that implements a psychophysical algorithm to estimate pain thresholds (QUEST Psychtoolbox, [23]). The algorithm incorporates the participant’s responses after each trial, yielding two psychometric functions that were fit using a Weibull distribution [22]. These functions represented the participant’s hot pain threshold when stimulation was applied and another psychometric function for when stimulation was off. This enables adaptation to the most likely pain threshold temperature. The full description of the psychophysical pain task and its capability is reported elsewhere [22].

### Patient Selection

Patients undergoing routine intracranial monitoring with sEEG for localization of epileptic foci were screened for inclusion in the study in 2022. A total of 4 patients (2 female, 2 male) met the inclusion criteria (see below) and were included in the final data analysis. The age range of the patient population was 21-40 years (mean 29 ± 8 years), and all patients had a diagnosis of drug-resistant epilepsy. All patients underwent sEEG for clinical purposes. Participation in the research study was obtained through informed consent under a protocol approved by the University of Utah Institutional Review Board.

To be eligible for the study, participants must have been 18 years of age or older, able to give informed consent, without any nerve damage in their arms or hands, able to communicate during the task, and not have any serious medical conditions such as bleeding disorders or cancer. Patients were excluded if their seizures interfered with data collection.

### Electrodes

The study utilized electrodes designed for sEEG. The locations of the electrodes were identified using each patient’s structural magnetic resonance imaging coregistered with postoperative computed tomography using the LeGUI software package (https://github.com/Rolston-Lab/LeGUI) [24]. The Brainnetome atlas was used within LeGUI to label the anatomical region based on the electrode locations (http://atlas.brainnetome.org) [25]. The atlas labels corresponding to each electrode were then converted to regional gyri by only keeping the first part of the alphanumeric atlas label. The distinction between the right and left hemispheres was also removed. For example, the Brainnetome label “CG_L_7_2” was collapsed to “CG.” Electrode locations were not included if they were within the white matter.

Patient specification and stimulation parameters are shown in **Table 1**. The electrodes sampling the cingulate brain regions were visualized in Montreal Neurological Institute (MNI) space using NeuroMArVL (https://github.com/mangstad/neuromarvl.

**Table 1.**
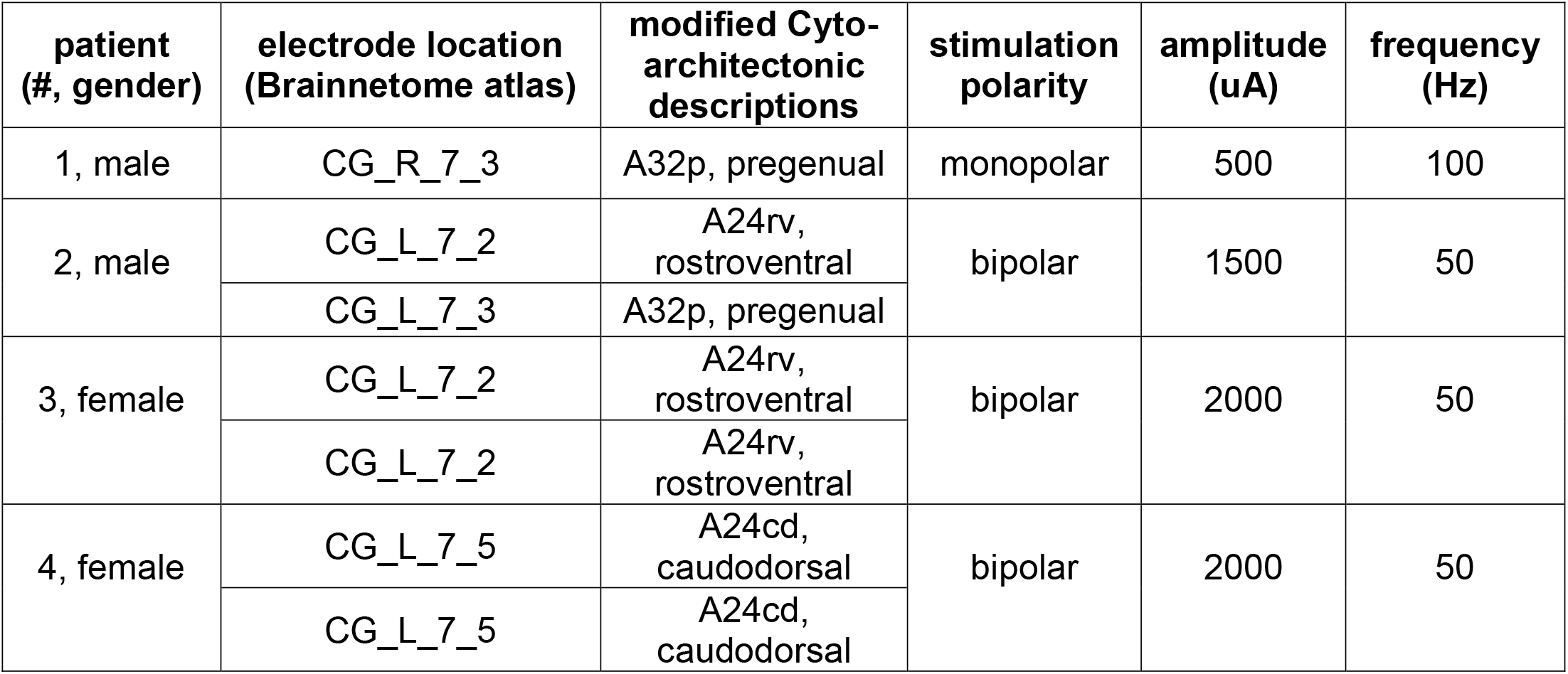
Stimulation parameters and location for all patients.

### Intracranial recording and stimulation

Neurophysiological data were recorded using a 128-channel data acquisition system (NeuroPort, Blackrock Microsystems, Salt Lake City, UT). Recorded data were bandpass filtered online at 0.3-250 Hz and sampled at 1 kHz. Reference channels were chosen for each patient consisting of a single intracranial depth electrode contact located in the white matter without artifact distal from the irritative zone. The reference electrode was selected after electrode location labeling. A disposable Bovie Grounding Pad was used as the return (or “ground”) electrode for stimulation to minimize noise and stimulation artifacts.

Stimulation was current controlled with biphasic rectangular pulses. We used bipolar and monopolar stimulation polarity, 500-2000 uA, 50 or 100 Hz, 2s train length, and a pulse width of 0.2 ms. If the stimulation was not well tolerated (i.e., caused discomfort to the patient or after-stimulation epileptiform discharges) or if the patient could reliably detect when stimulation was on, parameters such as polarity, amplitude, frequency, duration, or location were adjusted. Stimulation started when the capacitive touch sensor indicated that the patient put their hand on the thermoelectric device. Parameters were constrained following previous studies designed to avoid tissue injury, where the charge density per square pulse < 55 μC/cm^2^ [26,27]. Stimulation parameters for each patient were based on the pretesting monitoring procedure developed by Suthana et al. [28]. Stimulation was employed systematically with low limits to avoid causing patient discomfort. The research team and clinical staff continuously monitored the real-time intracranial recordings for patient safety.

All recruited patients had at least one contact within the cingulum for clinical purposes. We prioritized stimulating electrodes within Brodmann area 24, as a meta-analysis showed that the ACC, primarily area 24, participates in both the affective and attentional concomitants of pain sensation and response selection [29]. Area 32 is also considered part of the dACC. This region was considered if the patient did not have a contact in area 24. After selecting the stimulation location, we optimized stimulation parameters.

During intraoperative stimulation testing in the ventral posterolateral and ventral posteromedial nuclei, Boccard et al. found that DBS at frequencies ≤50 Hz were analgesic, but frequencies > 70 Hz were hyperalgesic in patients with neuropathic pain [13]. Liu et al. used both 50 Hz and 100 Hz in epilepsy patients undergoing pain threshold testing and found no significant difference between the two [30]. Therefore, our initial testing frequency was 50 Hz. Patient 1 described a tingling sensation with stimulation at low levels of stimulation (0.5 – 1mA) but did not describe this at 100 Hz and when the stimulation polarity was switched to monopolar. The remaining patients received stimulation at 50 Hz and bipolar stimulation.

Another parameter that varied between patients was the stimulation amplitude. We started safety testing at 0.5 mA and increased in steps of 0.5 mA. Stimulation was reduced in steps of 0.25 mA if the patient knew when stimulation was applied, or after-stimulation epileptiform discharges were observed. We were conservative in our stimulation limit of 2 mA, as limited data describes detailed safety limits of current controlled trains in the ACC.

Once the pretesting monitoring procedure was complete, the psychophysical pain task started. Stimulation was applied in a pseudorandom order to provide a uniform spatial and temporal charge distribution and avoid overstimulation of the area. The stimulation parameters and behavioral task were controlled using custom software written in Labview (National Instruments, Austin, TX).

### Preprocessing non-stimulation trials for mixed-effect modeling

We hypothesized that stimulation increased pain thresholds in patients with baseline neural encoding of pain psychophysics in the cingulate gyrus. We analyzed non-stimulation trials to investigate why some patients had shifted psychometric curves associated with stimulation and others did not. A 60 Hz notch filter was applied to remove line noise. Recordings were re-referenced using the common median, which is more robust to large amplitude transient events than the common average reference [31]. Data were segmented into epochs of -1000 ms to 1000 ms to facilitate analysis relative to specific trial events. These events included: 1) initiation of stimulus delivery upon participant’s hand placement on the device and 2) removal of the participant’s hand from the device. After removing their hand from the device, participants responded via a touchscreen monitor to whether the stimulus was painful. Participants subsequently provided a numerical rating of perceived pain intensity on a visual analog scale (VAS) of 0-10, which was not time-locked to the intracranial recordings. Both pain evaluation metrics were utilized as response variables in mixed-effects modeling.

Epileptiform discharges/noise spikes were identified as outliers if the epoch contained a value that was more than three median absolute deviations from the median. Channels were also visually inspected for artifacts after this automated analysis. Signals were downsampled to 500 Hz for subsequent analyses.

sEEG recordings allow evaluation of local neural activation through changes in amplitude within the broadband high-gamma frequency range (HFA; 70-150 Hz) in the recorded local field potential (LFP) [19–21]. HFA is highly correlated with both blood-oxygen-level-dependent fMRI and population firing rates, making it a useful tool for bridging the gap between human neuroimaging results and findings from animal model electrophysiology studies [32]. HFA of the cingulate cortex was isolated through bandpass filtering the LFP between 70-150 Hz using a 4th-order zero-phase Butterworth filter. The absolute value of the Hilbert transform was applied to isolate the amplitude of the HFA component. The signal was smoothed using a moving average filter with a 300 ms fixed window. The epochs for each channel were normalized against baseline activity, using the mean and standard deviation from -3 to -2 s relative to the trial event onset to represent the relevant event being modeled. The processing was similar to our prior work [33].

### Statistical analysis

Pain thresholds were compared using a left-tailed Welch’s T-test to test whether the pain threshold with stimulation was greater than the threshold without stimulation. We used sliding-window generalized linear mixed-effects (GLME) models to investigate the baseline relationship between the HFA component of the LFP within each patient’s cingulate cortex and the patient’s pain responses when stimulation was off. We focused our analyses on the first second the hand was exposed to the stimulus and the last second the hand was on the stimulus. We also evaluated the second after the hand was removed. We chose a time window of 500 ms with a 250 ms overlapping sliding window to capture the likely time course of physiological HFA of painful stimuli [34].

Two different types of GLME models were employed for each patient. The first modeled the relationship between HFA and whether the patient responded “yes” or “no” to whether the trial was painful (binary response). The fixed effect in this model was the mean z-scored HFA amplitude during the 500 ms window in all electrode contacts located in the cingulate cortex. The Wilkinson notation for this model was: y∼A+(A|B), where A was HFA amplitude, and B was the trial temperature on the thermoelectric device. A random intercept and slope model was specified to account for variation in the fixed effect across different temperatures. A binomial distribution was used because the response variable was binary.

The second model evaluated the relationship between HFA and the numerical pain rating on a scale from 0 to 10. As in the first model, the fixed effect was the mean HFA amplitude of the cingulate cortex during a 500 ms window. As previously, the random effect was trial temperature.

Statistical significance for each model was determined by a permutation test, where the model was re-evaluated 1,000 times with patients’ responses randomly permuted. The 95% range of the pseudo-t-statistics was used to determine the bounds of t-statistics associated with randomness. The true model was considered significant if the real t-statistic did not fall within these bounds. For the significantly predictive windows, an observed probability, *P*, was determined by adding the number of pseudo-t-statistics greater than the real t-statistic, multiplying by two to account for the two-sided distribution, and then dividing by the 1000 total observations. The smallest possible P-value was 0.002.

## Results

All four patients completed the psychophysical task with and without stimulation in the cingulate cortex. Participants completed at least 30 trials of each condition to allow adequate sampling for signal processing. Weibull probability density functions representing the likelihood that a temperature was painful were derived for each patient (**Figure 2**). The pain threshold was classified as the temperature with a 50% probability of that temperature being painful. All thresholds had a standard deviation of less than 0.5 ºC. Reported pain thresholds when stimulation was “on” compared to when it was “off” showed no difference for Patient 1 (t=2.5, p=0.98) and Patient 4 (t=1.3, p=0.89). However, Patient 2 and Patient 3 had a higher stimulation “on” pain threshold than when stimulation was “off.” The threshold when stimulation was off for Patient 2 was 53.74 ºC while it significantly increased to 54.16 ºC when stimulation was on (t=-4.9, p=0.0004). The threshold when stimulation was off for Patient 3 was 50.38 ºC and significantly increased to 50.72 ºC when stimulation was on (t=-3.5, p=0.002)

**Figure 2:**
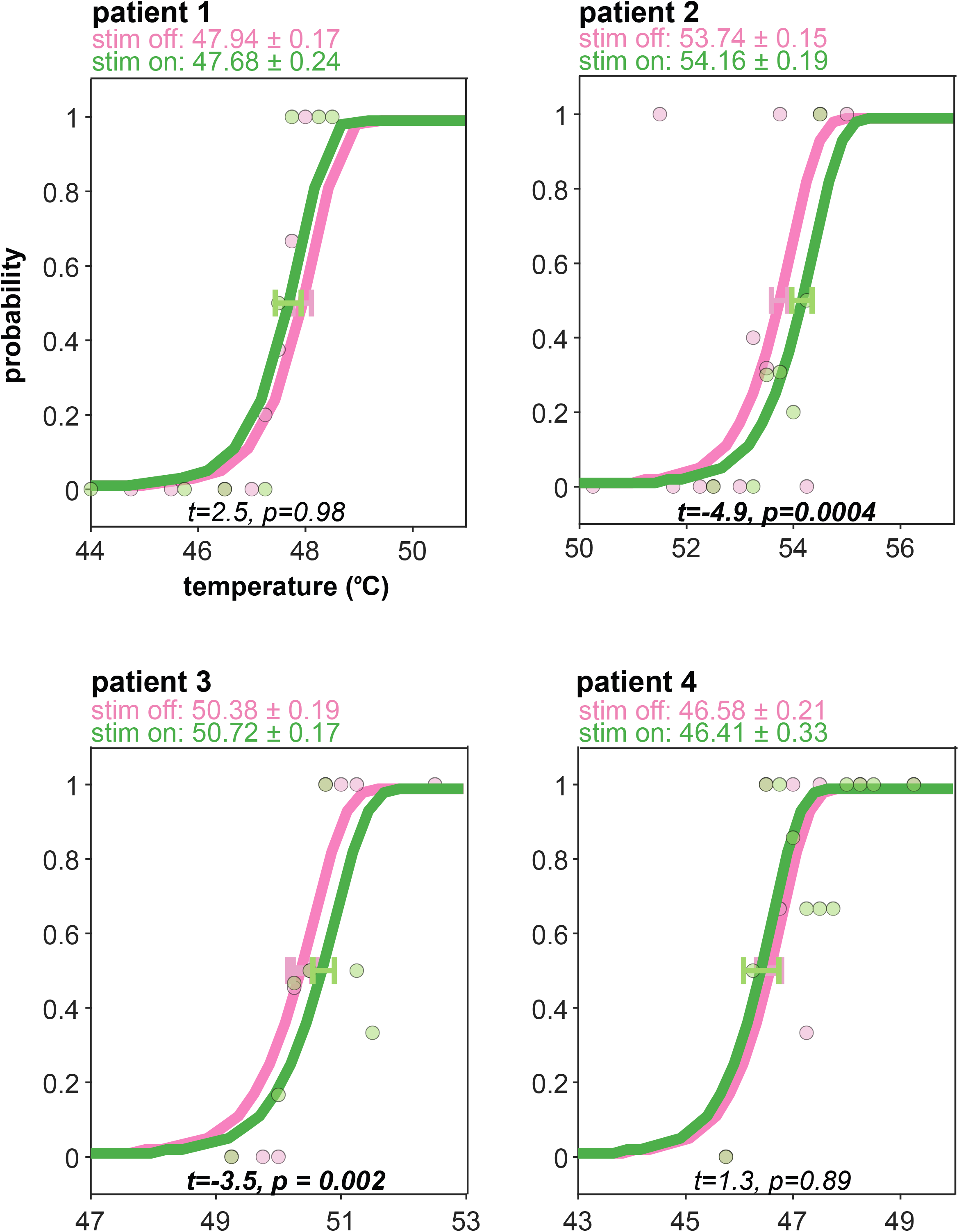
Psychophysical probability density functions of pain with and without stimulation. Shown are all four patients’ psychometric probability density functions. These curves represent the probability of pain at specific temperatures, and the circles represent the density of the probability based on experimental data. The pain threshold is the temperature with a 50% probability of the stimulus being painful. Each threshold is associated with a standard deviation, shown as a horizontal error on each curve. Patient 1 and Patient 4 showed no difference in pain thresholds when stimulation was on compared to off (Welch’s t-test). However, Patients 2 and 3 had a higher pain threshold with stimulation.

We hypothesized that increased hot pain thresholds with stimulation would occur in patients with psychophysical pain responses predicted by non-stimulation neural activity in their cingulate. The z-scored mean HFA amplitude during the first second of the stimulus, the last second of the stimulus, and the second after the stimulus ended were modeled (**Figure 3**). HFA in Patients 1 and 4 was neither predictive of the “yes/no” binary response variable nor the VAS ratings. However, HFA in Patient 2 predicted an increased VAS rating from -250 to 250 ms relative to the stimulus onset (P = 0.026). The t-statistic for both windows was positive, which can be interpreted as an increase in HFA was associated with an increase in VAS ratings. Patient 3 showed similar temporal predictability, as 0 ms to 500 ms after the stimulus had HFA that was predictive of whether they responded “no” to whether the trial was painful (P=0.036).

**Figure 3:**
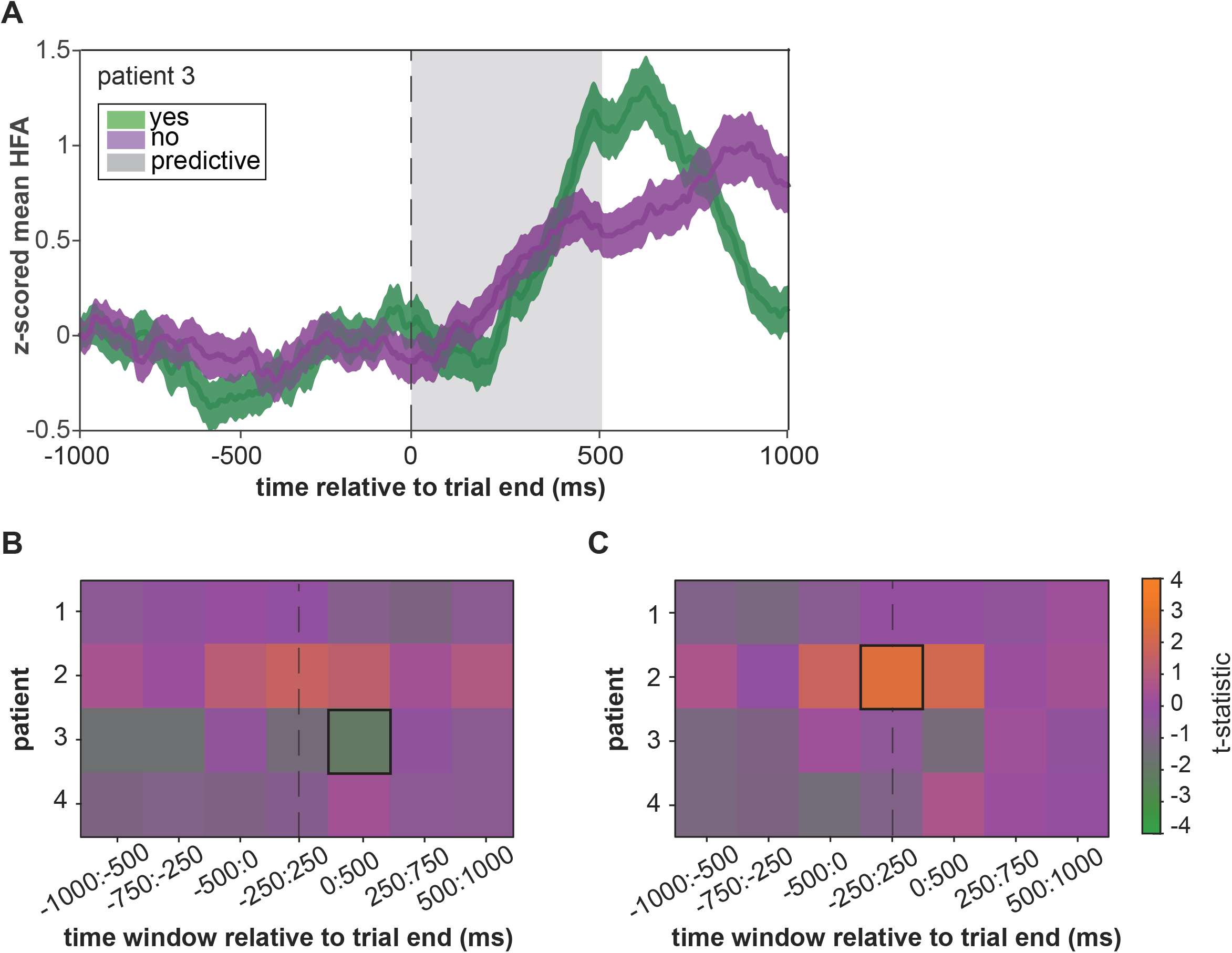
Relationship between HFA and the psychophysical evaluation of pain. 2A: The mean “yes” and “no” z-scored mean HFA for Patient 3 during the last second of the trial within the cingulate gyrus. The predictive time window from the heatmap in 2B is shaded in grey. It should be noted that visualizing the z-scored mean HFA across time does not account for interaction variables, such as temperature and patient identification even though the predictive regions did account for these variables through GLME modeling. 2B: Heatmap coloring represents the real t-statistics for the HFA in each patient’s cingulate gyrus. Each row in the heatmap represents one of the four patients. Each column represents one of the overlapping 500 ms windows that were analyzed. If the real t-statistic was significant (see *Statistical Analysis*), we outlined the region with a black box. The dotted vertical lines represent the end of the trial. Patient 3’s HFA predicted an increase in “no” responses from 0:500 ms after the stimulus ended (P=0.0360). 2C: Similar to 2B, except the response variable is the VAS score. Patient 2 had HFA predictive of an increase in VAS ratings from -250 ms to 250 ms relative to the end of the trial (P=0.0260)

## Discussion

The cingulate cortex, particularly the ACC, plays a crucial role in the affective and cognitive aspects of pain processing. Targeted neurosurgical pain treatment has been focused on the cingulate cortex, with recent use of deep brain stimulation (DBS) showing promising results. However, patient-specific factors can cause functional reorganization in neural connectivity over time, which must be considered when identifying structures and patients that will respond to DBS. We utilized intracranial neurophysiological recordings from four patients undergoing epilepsy surgery to assess the relationship between pain-related neural activity, behavioral responses, and pain threshold modulation using stimulation. Our findings suggest that patients who displayed increased pain-related neural activity responded to pain modulation through stimulation.

The first stereotactic cingulotomy for intractable pain was performed by Foltz and White [8], who reported that emotion factors significantly augmented incapacitating pain [7]. Recent case reports and series using standard DBS electrodes have shown mixed results [11,14,35]. For instance, Parvizi et al. explored the effects of stimulation of the dACC on two epilepsy patients with implanted intracranial electrodes. They found that stimulation led to cognitive, emotional, and autonomic reactions characterized as a “will to persevere” [10]. Resting-state fMRI suggested that this effect may result from activating networks that assign salience. Although the study investigated acute rather than chronic stimulation, it provides a framework to understand how electrical modulation of the dACC could produce affective pain relief by targeting the perceived emotional burden rather than the sensory experience itself. Bijanki et al. recently presented evidence to support the effectiveness of cingulum stimulation as an anxiolytic treatment. Specifically, they demonstrated a strong reduction in anxiety symptoms in three epilepsy patients who underwent sEEG [9]. Cingulotomies for intractable pain demonstrate the impact of emotional factors on pain perception. Exploration of ACC stimulation on cognitive and emotional aspects provides insight into the potential utility of affective pain relief through targeting perceived emotional burden.

Chronic pain patients are impacted by unique cognitive and emotional factors that can cause changes in functional neural networks [15]. Negative expectations can reverse the analgesic effect of remifentanil, while positive expectations are essential for placebo analgesia [36,37]. Distraction can also powerfully influence pain [38]. Pain catastrophizing can amplify pain intensity and unpleasantness in individuals with deficient coping strategies [39]. Fear-avoidance, maladaptive coping, and learning and memory are important factors in chronic pain [40]. Emotional states significantly influence pain—a negative emotional state can increase pain, while a positive state can lower it [41]. Empathy for others can even increase pain [42]. Although the direction of causality is difficult to determine, several recent studies have shown that structural and functional changes are reversible after chronic pain is resolved, suggesting that chronic pain leads to these changes [43–46]. Historical cingulotomies and successful case studies of cingulum DBS suggest the reverse is also true. Disrupting the functionality of the cingulum can reduce pain in some patients.

A larger case series (n=11) that tried dACC DBS did not produce as positive results [12]. Although patients with various chronic pain conditions initially reported improvements in social function, bodily pain, and overall score on the EQ-5D, only a few patients still had effective pain relief at the three-year follow-up [14]. Overall outcome measures returned to baseline. Despite patients reporting that their pain was present, they described it as “less bothersome” or “separate from them,” underscoring the affective role of the ACC [47]. We believe that larger DBS trials for pain have not been successful because a one-size-fits-all model is not appropriate, and a different approach is needed [48,49].

Individualized assessments of neural activity may be necessary to develop targets for stimulation. We hypothesized that pain thresholds would increase with cingulate stimulation in patients participating in our psychophysical pain experiment. After only seeing an effect from stimulation in half of the patients, we evaluated the baseline activity in all the patients to see if HFA in the cingulate was predictive, on an individual patient basis, in the patient’s psychophysical evaluation of pain. Interestingly, the patients who had HFA amplitudes related to their pain response at some point at the onset and end of the stimulus were the same patients who responded to stimulation. HFA was related to a decrease in psychophysical pain in Patient 2 but an increase in psychophysical pain in Patient 3. This contrasting result may be related to different pain coping mechanisms in the two patients. While we acknowledge this finding needs to be substantiated with more patients and electrode locations, we propose that areas involved in the psychophysical evaluation of pain may be good biomarkers in understanding where to target stimulation.

There were a few limitations in addition to the small sample size of the case series. For safety reasons, the investigator conducting the study was not blinded to when stimulation was applied, and the patient was constantly monitored for signs of overstimulation electrographically and clinically. The stimulation parameters were adjusted to be undetectable by the patient, which may have resulted in a smaller effect or sub-behavioral effect. Additionally, the stimulation electrode varied slightly in all four patients due to the clinical indication for sEEG being seizure localization. Patients 2 and 3 had contacts closer to the rostroventral cingulate. Patient 1 had their stimulation electrode in the pregenual cingulate, and Patient 4’s electrodes were in the caudodorsal cingulate. We also used a variety of stimulation parameters to maximize the effect while accounting for patient safety. This is a realistic picture of how DBS is used in a clinical setting, as patients may not tolerate high stimulation levels and adjustments may be necessary.

## Conclusion

We report on four patients who underwent electrical stimulation of the cingulate cortex while performing a pain task. Our hypothesis was that this stimulation would raise the pain threshold and we observed this effect in certain patients. To investigate why the stimulation appeared to alter pain perception in some patients, we examined the HFA associated with pain during baseline psychophysical evaluations. Our results indicated that patients who exhibited HFA during pain perception without stimulation were the same ones who responded to the stimulation. Therefore, personalized evaluation of neural activity biomarkers could be beneficial in pinpointing the optimal target for stimulation and predicting its effectiveness.

## Data Availability

All data produced in the present study are available upon reasonable request to the authors

## Acknowledgements

Thank you to Greg Stoddard for providing statistical consulting.

## References

[1] Tan LL, Kuner R. Neocortical circuits in pain and pain relief. Nat Rev Neurosci 2021:1–14. https://doi.org/10.1038/s41583-021-00468-2.

[2] Etkin A, Egner T, Kalisch R. Emotional processing in anterior cingulate and medial prefrontal cortex. Trends Cogn Sci 2011;15:85–93. https://doi.org/10.1016/j.tics.2010.11.004.

[3] Kucyi A, Davis KD. The dynamic pain connectome. Trends Neurosci 2015;38:86–95. https://doi.org/10.1016/j.tins.2014.11.006.

[4] Caston RM, Smith EH, Davis TS, Rolston JD. The Cerebral Localization of Pain: Anatomical and Functional Considerations for Targeted Electrical Therapies. J Clin Med 2020;9:1945. https://doi.org/10.3390/jcm9061945.

[5] Shackman AJ, Salomons T v., Slagter HA, Fox AS, Winter JJ, Davidson RJ. The integration of negative affect, pain and cognitive control in the cingulate cortex. Nat Rev Neurosci 2011;12:154–67. https://doi.org/10.1038/nrn2994.

[6] Russo JF, Sheth SA. Deep brain stimulation of the dorsal anterior cingulate cortex for the treatment of chronic neuropathic pain. Neurosurg Focus 2015;38:E11. https://doi.org/10.3171/2015.3.FOCUS1543.

[7] Foltz EL, White LE. The role of rostral cingulumotomy in “pain” relief. Int J Neurol 1968;6:353–73.

[8] Foltz EL, White LE. Pain “Relief” by Frontal Cingulumotomy. J Neurosurg 1962;19:89–100. https://doi.org/10.3171/jns.1962.19.2.0089.

[9] Bijanki KR, Manns JR, Inman CS, Choi KS, Harati S, Pedersen NP, et al. Cingulum stimulation enhances positive affect and anxiolysis to facilitate awake craniotomy. Journal of Clinical Investigation 2019;129:1152–66. https://doi.org/10.1172/JCI120110.

[10] Parvizi J, Rangarajan V, Shirer W, Desai N, Greicius MD. The Will to Persevere Induced by Electrical Stimulation of the Human Cingulate Gyrus. Neuron 2013;80. https://doi.org/10.1016/j.neuron.2013.10.057.The.

[11] Spooner J, Yu H, Kao C, Sillay K, Konrad P. Neuromodulation of the cingulum for neuropathic pain after spinal cord injury. Case report. J Neurosurg 2007;107:169–72. https://doi.org/10.3171/JNS-07/07/0169.

[12] Boccard SGJ, Fitzgerald JJ, Pereira EAC, Moir L, van Hartevelt TJ, Kringelbach ML, et al. Targeting the Affective Component of Chronic Pain. Neurosurgery 2014;74:628–37. https://doi.org/10.1227/NEU.0000000000000321.

[13] Boccard SGJ, Pereira EAC, Moir L, Aziz TZ, Green AL. Long-term outcomes of deep brain stimulation for neuropathic pain. Neurosurgery 2013;72:221–30. https://doi.org/10.1227/NEU.0b013e31827b97d6.

[14] Boccard SGJ, Prangnell SJ, Pycroft L, Cheeran B, Moir L, Pereira EAC, et al. Long-Term Results of Deep Brain Stimulation of the Anterior Cingulate Cortex for Neuropathic Pain. World Neurosurg 2017;106:625–37. https://doi.org/10.1016/j.wneu.2017.06.173.

[15] Zheng W, Woo CW, Yao Z, Goldstein P, Atlas LY, Roy M, et al. Pain-Evoked Reorganization in Functional Brain Networks. Cerebral Cortex 2020;30:2804–22. https://doi.org/10.1093/cercor/bhz276.

[16] Markman T, Liu CC, Chien JH, Crone NE, Zhang J, Lenz FA. EEG analysis reveals widespread directed functional interactions related to a painful cutaneous laser stimulus. J Neurophysiol 2013;110:2440–9. https://doi.org/10.1152/jn.00246.2013.

[17] Kim JH, Chien JH, Liu CC, Lenz FA. Painful cutaneous laser stimuli induce eventrelated gamma-band activity in the lateral thalamus of humans. J Neurophysiol 2015;113:1564–73. https://doi.org/10.1152/jn.00778.2014.

[18] Liu CC, Chien JH, Chang YW, Kim JH, Anderson WS, Lenz FA. Functional Role of Induced Gammy Oscillatory Responses in Processing Noxious and Innocuous Sensory Events in Humans. Neuroscience 2015;310:389–400. https://doi.org/10.1016/j.neuroscience.2015.09.047.

[19] Ray S, Maunsell JHR. Different origins of gamma rhythm and high-gamma activity in macaque visual cortex. PLoS Biol 2011;9. https://doi.org/10.1371/journal.pbio.1000610.

[20] Bartoli E, Bosking W, Chen Y, Li Y, Sheth SA, Beauchamp MS, et al. Functionally Distinct Gamma Range Activity Revealed by Stimulus Tuning in Human Visual Cortex. Current Biology 2019;29:3345–3358.e7. https://doi.org/10.1016/j.cub.2019.08.004.

[21] Buzsáki G, Anastassiou CA, Koch C. The origin of extracellular fields and currents--EEG, ECoG, LFP and spikes. Nat Rev Neurosci 2012;13:407–20. https://doi.org/10.1038/nrn3241.

[22] Caston RM, Davis TS, Smith EH, Rahimpour S, Rolston D. A novel thermoelectric device integrated with a psychophysical paradigm to study pain processing in human subjects. J Neurosci Methods 2023;386:109780. https://doi.org/10.1016/j.jneumeth.2022.109780.

[23] Watson AB, Pelli DG. QUEST: A Bayesian adaptive psychometric method. vol. 33. 1983.

[24] Davis TS, Caston RM, Philip B, Charlebois CM, Anderson DN, Weaver KE, et al. LeGUI: A Fast and Accurate Graphical User Interface for Automated Detection and Anatomical Localization of Intracranial Electrodes. Front Neurosci 2021;15. https://doi.org/10.3389/fnins.2021.769872.

[25] Fan L, Li H, Zhuo J, Zhang Y, Wang J, Chen L, et al. The Human Brainnetome Atlas: A New Brain Atlas Based on Connectional Architecture. Cereb Cortex 2016;26:3508–26. https://doi.org/10.1093/cercor/bhw157.

[26] Gordon B, Lesser RP, Rance NE, Hart J, Webber R, Uematsu S, et al. Parameters for direct cortical electrical stimulation in the human: histopathologic confirmation. Electroencephalogr Clin Neurophysiol 1990. https://doi.org/10.1016/0013-4694(90)90082-U.

[27] Ostrowsky K, Magnin M, Ryvlin P, Isnard J, Guenot M, Mauguière F. Representation of Pain and Somatic Sensation in the Human Insula: a Study of Responses to Direct Electrical Cortical Stimulation. Cerebral Cortex 2002;12:376–85. https://doi.org/10.1093/cercor/12.4.376.

[28] Suthana N, Haneef Z, Stern J, Mukamel R, Behnke E, Knowlton B, et al. Memory enhancement and deep-brain stimulation of the entorhinal area. New England Journal of Medicine 2012. https://doi.org/10.1056/NEJMoa1107212.

[29] Peyron R, Laurent B, Garcia-Larrea L. Functional imaging of brain responses to pain. Neurophysiol Clin 2000;30:263–88.

[30] Liu C-C, Moosa S, Quigg M, Elias WJ. Anterior insula stimulation increases pain threshold in humans: a pilot study. J Neurosurg 2021:1–6. https://doi.org/10.3171/2020.10.jns203323.

[31] Rolston JD, Gross RE, Potter SM. Common median referencing for improved action potential detection with multielectrode arrays. 2009 Annual International Conference of the IEEE Engineering in Medicine and Biology Society, IEEE; 2009, p. 1604–7. https://doi.org/10.1109/IEMBS.2009.5333230.

[32] Miller KJ. Broadband spectral change: Evidence for a macroscale correlate of population firing rate? Journal of Neuroscience 2010. https://doi.org/10.1523/JNEUROSCI.6401-09.2010.

[33] Caston RM, Smith EH, Davis TS, Singh H, Rahimpour S, Rolston JD. Characterization of spatiotemporal dynamics of binary and graded tonic pain in humans using intracranial recordings. BioRxiv 2023:2023.03.08.531576. https://doi.org/10.1101/2023.03.08.531576.

[34] Bastuji H, Frot M, Perchet C, Magnin M, Garcia-Larrea L. Pain networks from the inside: Spatiotemporal analysis of brain responses leading from nociception to conscious perception. Hum Brain Mapp 2016. https://doi.org/10.1002/hbm.23310.

[35] Boccard S, Pereira E, Moir L, Hartevelt T van, Kringelbach M, FitzGerald J, et al. Deep brain stimulation of the anterior cingulate cortex. Neuroreport 2014;25:83–8. https://doi.org/10.1097/WNR.0000000000000039.

[36] Bingel U, Wanigasekera V, Wiech K, Ni Mhuircheartaigh R, Lee MC, Ploner M, et al. The effect of treatment expectation on drug efficacy: imaging the analgesic benefit of the opioid remifentanil. Sci Transl Med 2011;3:70ra14. https://doi.org/10.1126/scitranslmed.3001244.

[37] Benedetti F, Mayberg HS, Wager TD, Stohler CS, Zubieta J-K. Neurobiological mechanisms of the placebo effect. J Neurosci 2005;25:10390–402. https://doi.org/10.1523/JNEUROSCI.3458-05.2005.

[38] Villemure C, Bushnell CM. Cognitive modulation of pain: how do attention and emotion influence pain processing? Pain 2002;95:195–9. https://doi.org/10.1016/S0304-3959(02)00007-6.

[39] Wade JB, Riddle DL, Price DD, Dumenci L. Role of pain catastrophizing during pain processing in a cohort of patients with chronic and severe arthritic knee pain. Pain 2011;152:314–9. https://doi.org/10.1016/j.pain.2010.10.034.

[40] Hashmi JA, Baliki MN, Huang L, Baria AT, Torbey S, Hermann KM, et al. Shape shifting pain: Chronification of back pain shifts brain representation from nociceptive to emotional circuits. Brain 2013;136:2751–68. https://doi.org/10.1093/brain/awt211.

[41] Villemure C, Bushnell MC. Mood influences supraspinal pain processing separately from attention. J Neurosci 2009;29:705–15. https://doi.org/10.1523/JNEUROSCI.3822-08.2009.

[42] Loggia ML, Mogil JS, Bushnell MC. Empathy hurts: compassion for another increases both sensory and affective components of pain perception. Pain 2008;136:168–76. https://doi.org/10.1016/j.pain.2007.07.017.

[43] Gwilym SE, Filippini N, Douaud G, Carr AJ, Tracey I. Thalamic atrophy associated with painful osteoarthritis of the hip is reversible after arthroplasty: A longitudinal voxel-based morphometric study. Arthritis Rheum 2010;62:2930–40. https://doi.org/10.1002/art.27585.

[44] Obermann M, Nebel K, Schumann C, Holle D, Gizewski ER, Maschke M, et al. Gray matter changes related to chronic posttraumatic headache. Neurology 2009;73:978–83. https://doi.org/10.1212/WNL.0b013e3181b8791a.

[45] Seminowicz DA, Wideman TH, Naso L, Hatami-Khoroushahi Z, Fallatah S, Ware MA, et al. Effective treatment of chronic low back pain in humans reverses abnormal brain anatomy and function. Journal of Neuroscience 2011;31:7540–50. https://doi.org/10.1523/JNEUROSCI.5280-10.2011.

[46] Rodriguez-Raecke R, Niemeier A, Ihle K, Ruether W, May A. Brain gray matter decrease in chronic pain is the consequence and not the cause of pain. Journal of Neuroscience 2009;29:13746–50. https://doi.org/10.1523/JNEUROSCI.3687-09.2009.

[47] Farrell SM, Green A, Aziz T. The Current State of Deep Brain Stimulation for Chronic Pain and Its Context in Other Forms of Neuromodulation. Brain Sci 2018;8:158. https://doi.org/10.3390/brainsci8080158.

[48] Shirvalkar P, Veuthey TL, Dawes HE, Chang EF. Closed-loop deep brain stimulation for refractory chronic pain. Front Comput Neurosci 2018;12. https://doi.org/10.3389/fncom.2018.00018.

[49] Mackey S, Greely HT, Martucci KT. Neuroimaging-based pain biomarkers: Definitions, clinical and research applications, and evaluation frameworks to achieve personalized pain medicine. Pain Rep 2019;4:1–13. https://doi.org/10.1097/PR9.0000000000000762.

